# Local genetic covariance between serum urate and kidney function obtained from local Bayesian regressions

**DOI:** 10.1101/2021.03.31.21254729

**Authors:** Alexa S Lupi, Nicholas A Sumpter, Megan P Leask, Justin O’Sullivan, Tayaza Fadason, Gustavo de los Campos, Tony R Merriman, Richard J Reynolds, Ana I Vazquez

## Abstract

Hyperuricemia is associated with several cardiometabolic and renal diseases, such as gout and chronic kidney disease. Previous studies have examined the shared genetic basis of chronic kidney disease and hyperuricemia either using single-variant tests or estimating whole-genome genetic correlations between the traits. Individual variants typically explain a small fraction of the genetic correlation between traits, thus reducing the power to map pleiotropic loci. Alternatively, genome-wide estimates of genetic correlation, while useful, do not shed light on what regions may be implicated in the shared genetic basis of traits. Therefore, to fill the gap between these two approaches, we used local Bayesian regressions to estimate the genetic covariance between markers for chronic kidney disease and hyperuricemia in specific genomic regions. We identified 267 linkage disequilibrium segments with statistically significant covariance estimates, 17 of which had a positive directionality and 250 negative, the latter being consistent with the directionality of the overall genetic covariance. These 267 significant segments implicated 188 genetically distinct shared loci. Many of these loci validate previously identified shared loci with consistent directionality, including 22 loci previously identified as shared. Numerous novel shared loci were also identified, such as *THBS3/MTX1/GBAP1, LINC01101, SLC7A9/CEP89, CYP24A1, KCNS3, CHD9, ARL15, PAX8*, and *IGF1R*. Finally, to examine potential biological mechanisms for these shared loci, we have implicated a subset of the genomic segments that are associated with gene expression using colocalization analyses. In particular, five genes (*FGF5, ARL6IP5, TRIM6, BCL2L1*, and *NTRK1*) expressed in the kidney are causal candidates potentially contributing to pleiotropic pathways between chronic kidney disease and hyperuricemia. The regions identified by our local Bayesian regression approach may help untangle and explain the association between chronic kidney disease and hyperuricemia.

**Author Summary:** Chronic kidney disease is of increased prevalence among people with hyperuricemia, suggesting a shared genetic etiology. Since markers for chronic kidney disease and hyperuricemia have an overall non-zero genetic correlation, there appears to be genetic basis to the shared etiology. However, genome-wide genetic correlation estimates do not elucidate the specific genomic regions contributing to both traits, particularly regions that contribute to the traits with opposite directionality to the overall directionality. We have implemented local Bayesian regressions to identify small genomic segments contributing to the overall genetic correlation. Our method is applicable to any pair of traits that have a shared genetic relationship. We have found numerous novel shared loci, validated previously reported loci, and identified new shared pathways simultaneously contributing to the markers between chronic kidney disease and hyperuricemia. These loci all merit detailed investigation as they may involve underlying biological mechanisms with the potential to explain the common pathogenesis of hyperuricemia and chronic kidney disease.

## Introduction

Chronic kidney disease (CKD) carries a significant global health and economic burden [1,2]. In the United States alone, it is estimated that 37 million adults (∼15%) have CKD and kidney diseases are the ninth leading cause of death [3]. CKD stages 3-5 manifest as decreased renal function and are defined by elevated serum creatinine (sCr) or estimated glomerular filtration rate (eGFR) <60 mL/min/1.73m^2^. CKD can lead to lower quality of life, increased risk of cardiovascular morbidity, and premature mortality [2]. Hyperuricemia is defined by serum urate (sU) concentration >6.8 mg/dL and is contributed to by deteriorating renal function [3]. Hyperuricemia has several comorbidities associated with it, including CKD [4], and can result in monosodium urate crystal deposits in joints and tendons, which leads to the development of gout. In the United States, an estimated 9.2 million people have gout (∼ 4%), which is also associated with substantial cardiovascular morbidity and all-cause mortality [5–8]. Among people with hyperuricemia there is a higher prevalence of CKD, and among patients with CKD, sU concentrations are higher [9,10].

Genome-wide analyses have demonstrated that the association observed between eGFR and serum urate has a genetic basis. Tin *et al*. carried out a large-sample trans-ethnic genome-wide association study (GWAS) of sU and, through cross-trait linkage disequilibrium (LD) score regression, obtained an estimate of overall genetic correlation between eGFR and sU of -0.26 (standard error of 0.04) [11]. This was one of the largest negative correlations with sU out of 748 traits analysed [11]. Reynolds *et al*., using two large family-based datasets and Bayesian whole-genome regressions, obtained global genetic correlations between sCr (which has a direct inverse relationship to eGFR, hence the directionality difference between the estimates) and sU of 0.20 (95% confidence interval (CI): 0.07, 0.33) in one dataset and 0.25 (95% CI: 0.07, 0.41) in the other [12]. However, the pleiotropic regions of the genome and biological mechanisms underlying the genetic relationship are unclear without identifying local genetic covariances [13].

GWAS of sU and eGFR have identified numerous loci associated with each phenotype separately. A recent study comparing large GWAS of the markers identified 35 shared loci [14]. However, the GWAS methods used to detect the shared signals used single-marker regressions or tests, which are based on the marginal association of individual single-nucleotide polymorphisms (SNPs) with phenotypes and thus do not account for LD between SNPs. Our method improves over post-analysis of GWAS summary statistics by estimating neighbouring SNP effects concomitantly. Incorporating local LD to estimate genetic effects in a tightly segregating chromosomal segment has been previously suggested [15–17].

In this study, we mapped the shared genetic basis of eGFR and sU using local Bayesian regressions (LBR) that estimate local genetic variances and covariances and capture LD patterns [17]. Our aim was to characterize the common genetic basis for CKD (eGFR) and hyperuricemia (serum urate levels) to disentangle the relationship through the identification and preliminary examination of pleiotropic genomic regions. We estimated local genetic covariances between sU and eGFR genome wide. We identified numerous local genetic regions as significant for local genetic covariance, including previously implicated shared loci and novel shared loci.

## Results

The study was based on the UK Biobank dataset and included 333,542 distantly related white participants, of whom 53.7% were female with an average age of 56.9 ± 8.0 years old. The average sCr level was 0.8 ± 0.2 mg/dL (the average ± standard error), eGFR was 144.2 ± 56.0 ml/min/1.73 m^2^, and the average sU level was 5.2 ± 1.3 mg/dL. Two (2.0) percent of the individuals had an ICD10 diagnosis or self-diagnosis of gout, 12.4% had hyperuricemia, 0.5% had CKD, and 0.3% had hyperuricemia and CKD. Our genetic analyses utilized directly genotyped autosomal SNPs from the UK Biobank Axiom™ Array by Affymetrix. After applying filters for minor-allele frequency ≥ 1% and for a missing call rate ≥ 5%, a total of 607,490 SNPs were used.

We identified 511,828 overlapping LD segments (small, non-independent chromosomal segments). Following Funkhouser *et al*. [17], we analysed the markers using a sequence of LBR, where each marker is regressed on contiguous SNPs in a large chromosomal segment plus overlapping flanking buffers (represented in S1 Fig). We collected the samples from the posterior distribution of effects for each LBR and used these samples to estimate the local variances for each marker (Fig 1) and the local covariances between the markers (Fig 2). Variances and covariances were computed within 511,828 LD segments identified. The LBRs were implemented using the BGLR R package [18], and had a variable selection prior distribution for the SNP effects with a point of mass at zero. A detailed description is provided in the Materials and Methods section.

**Fig 1.**
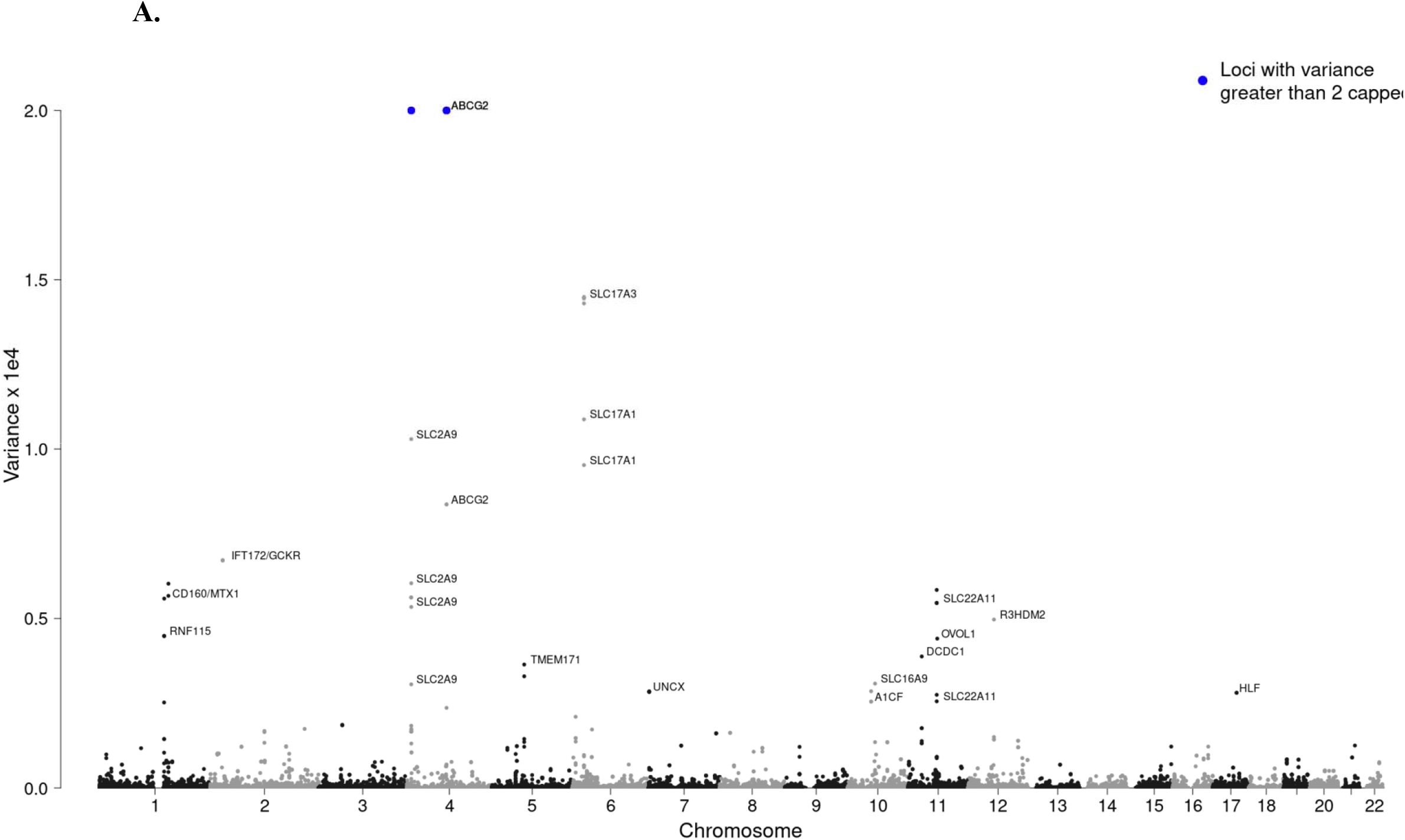
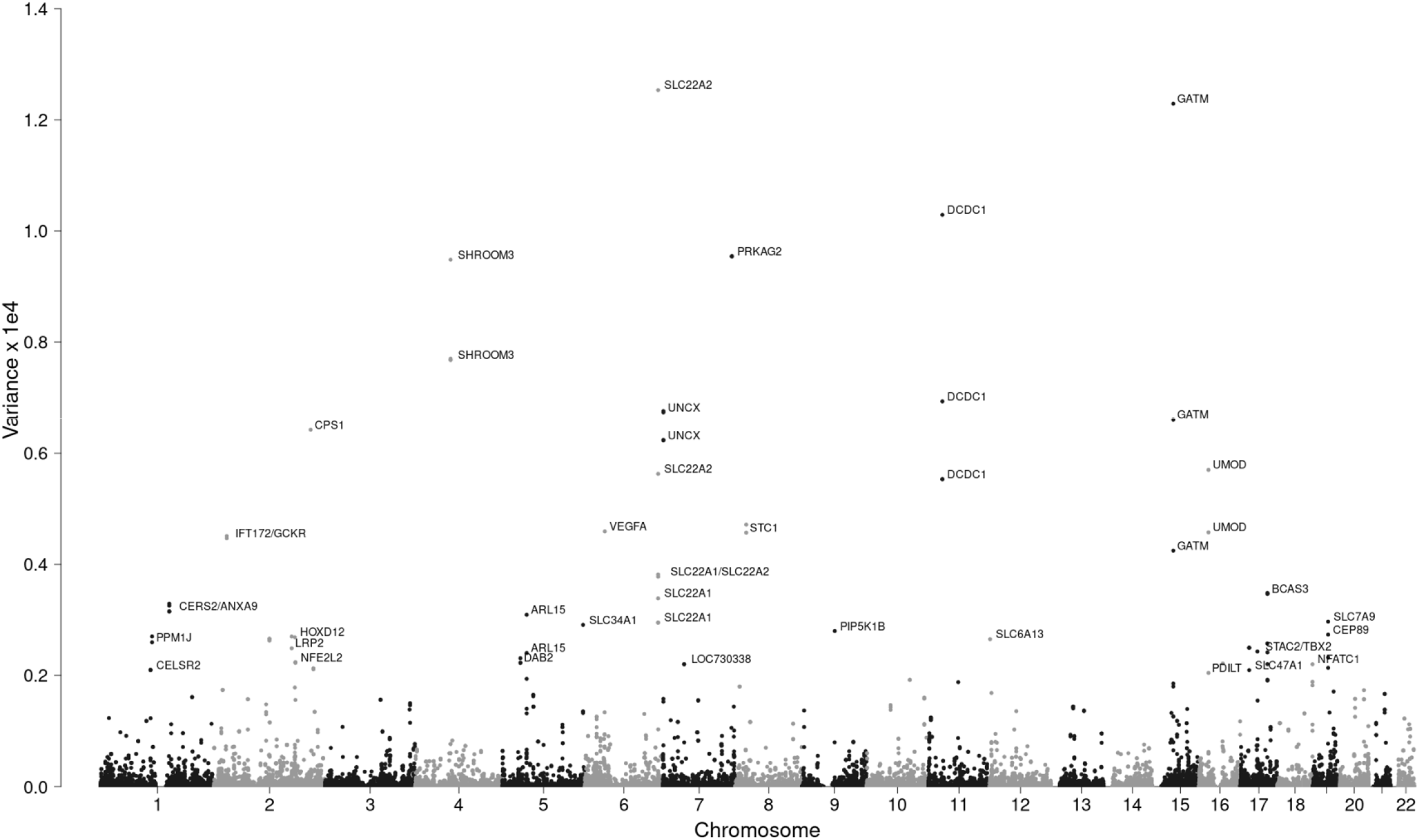
The variance estimates of LD segments in the unrelated white cohort of the UK Biobank for sU concentrations (A) and eGFR (B).

**Fig 2.**
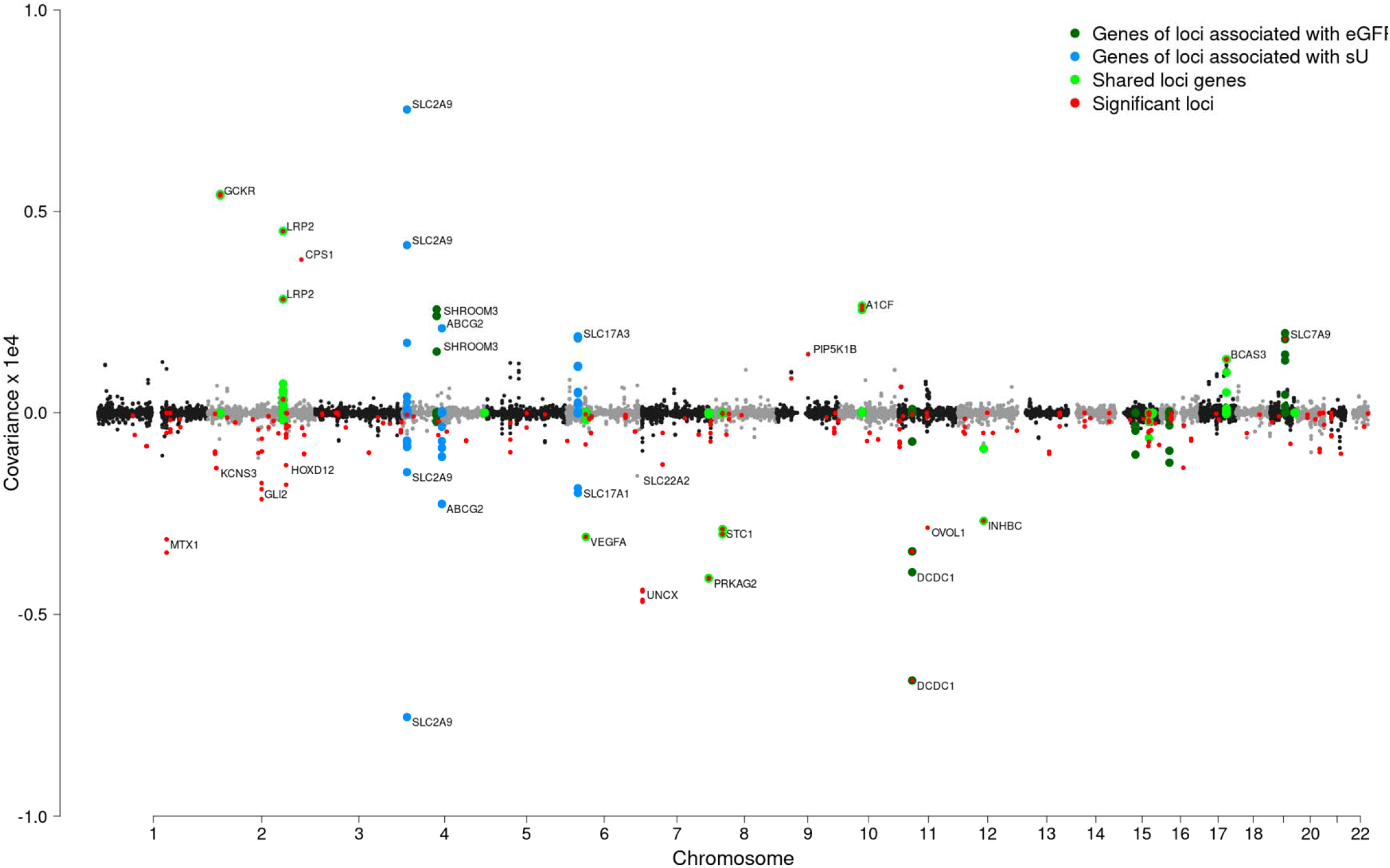
Covariance estimates of LD segments in UK Biobank, selectively annotated with the gene name of the mid-point SNP of that segment and the segment size. Segments that contained SNPs in loci associated with known eGFR genes are highlighted in dark green, segments that contained SNPs in genes associated with sU are highlighted in blue, and segments that contained SNPs in genes associated with both sU and eGFR (from comparing separate GWAS, Johnson et al. [21]) are highlighted in lime green. Segments significant for genetic covariance are highlighted in red.

Using a bootstrap resampling method, we obtained standard error estimates of the local genetic covariance estimates and found 267 LD segments where the covariance estimates had a 95% CI that did not include zero (Fig 2; S1 Table). Due to the computational burden of bootstrapping with a very large sample size, we preselected large genomic regions for bootstrapping if at least one SNP from a single-marker regression was significant for either sU or for a CKD marker (see methods for details and S2 Table for GWAS results). The number of SNPs in the significant LD segments ranged from one to 17, averaging 4.1 per segment (about 0.02 MB, excluding the 87 single SNP segments). Interestingly, 17 of the 267 significant segments showed positive genetic covariance estimate directionality, and the remaining 250 were negative estimates. After a conservative Bonferroni correction for multiple testing (see materials and methods section), 18 segments were still significant (S1 Table).

The 267 significant LD segments often included the same variants and map to identical GWAS loci, so we collapsed these 267 segments to 188 unique loci that possess genetic covariance signal between eGFR and sU (S3 Table). The top distinct loci implicated by the significant segments in terms of covariance magnitude are listed in Table 1. A graphical representation of some of the top significant loci, *i*.*e*., the top covariance magnitudes in significant distinct loci, is presented in Fig 3.

**Table 1.**
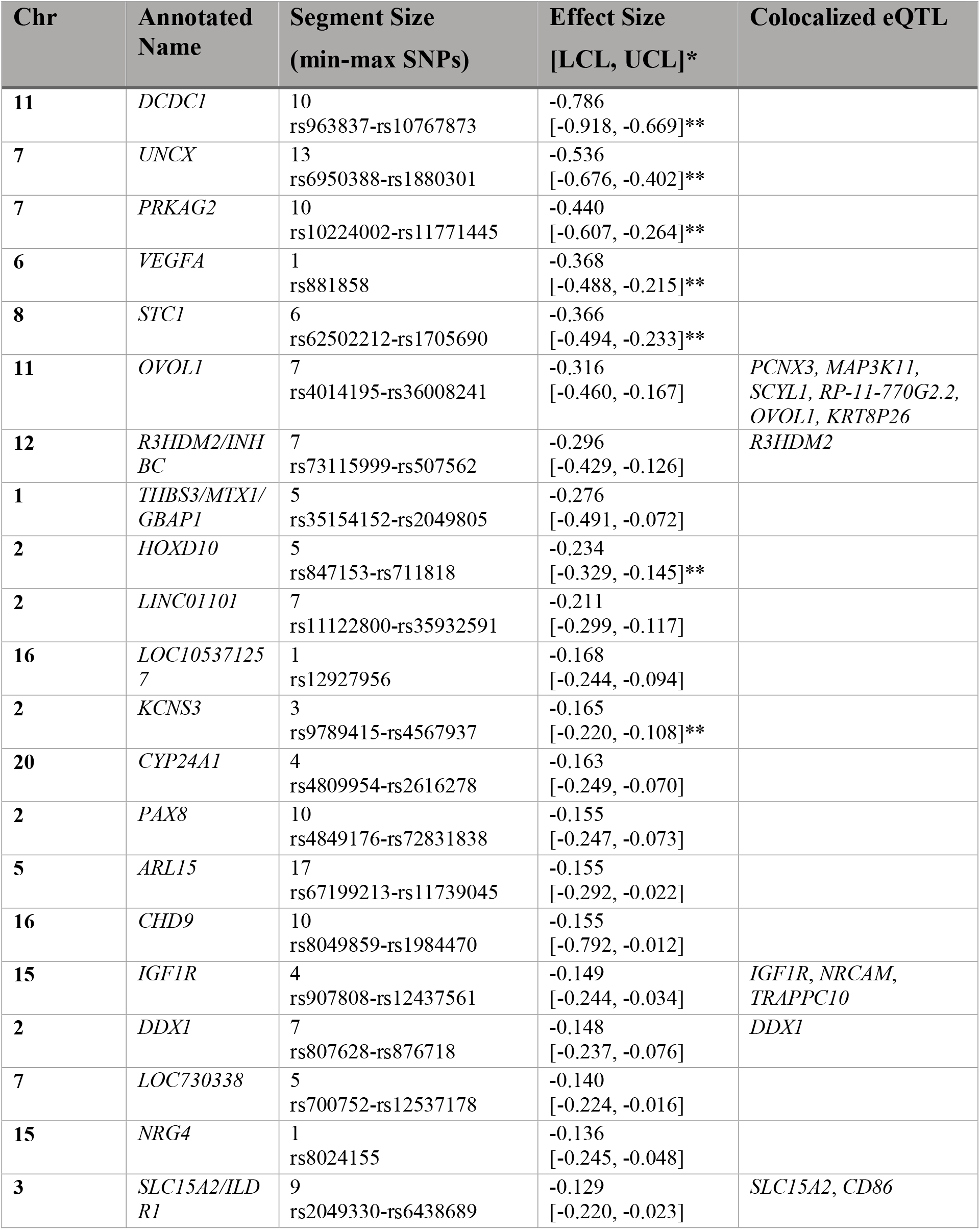
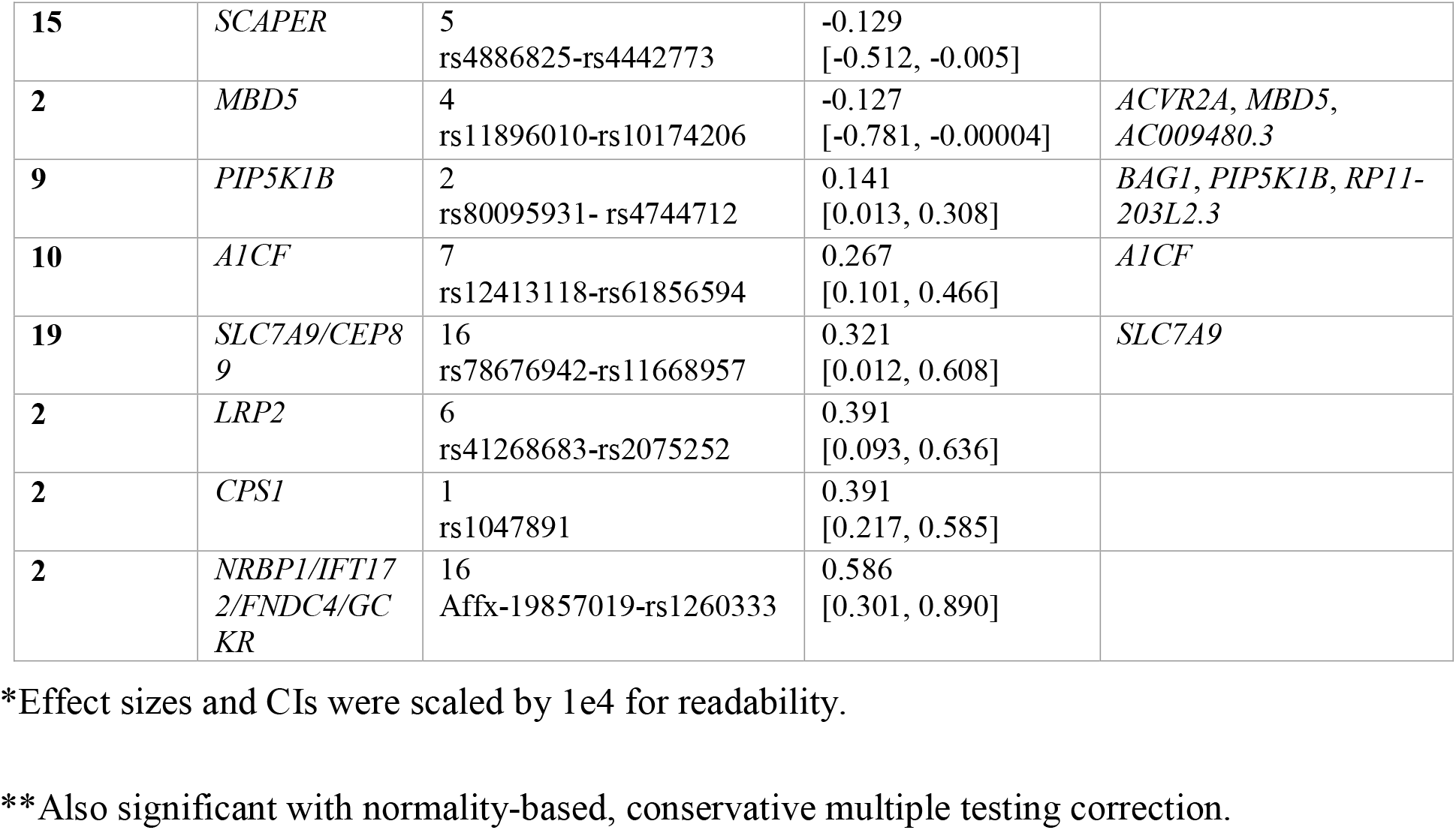
The top magnitude genomic segments significant for covariance between sU and eGFR with their chromosome, annotated name, effect size [95% CI], and colocalized eQTL.

**Fig 3.**
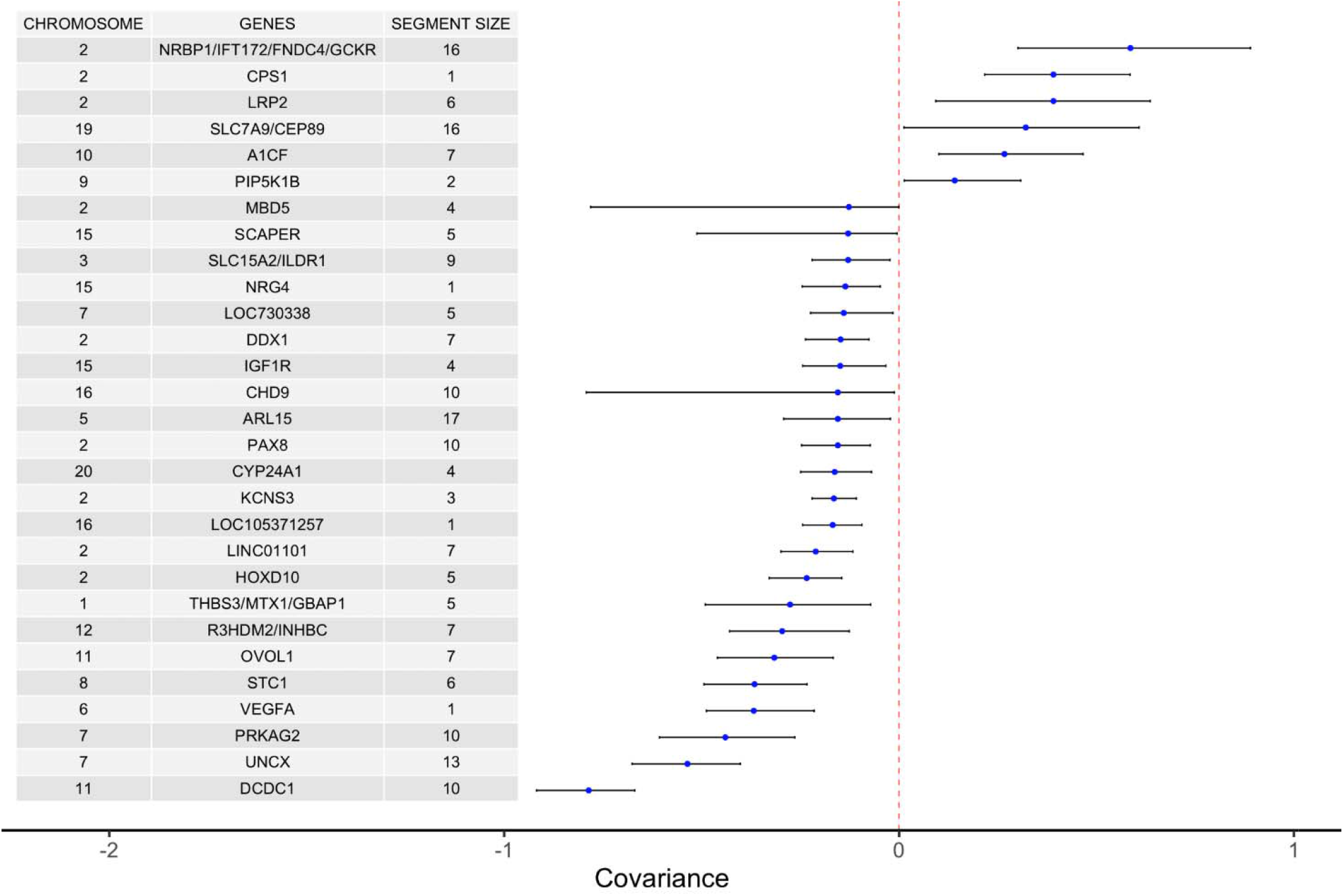
Figure of top 29 implicated significant shared loci (the distinct loci derived from the top 50 results) and their effects with corresponding 95% confidence intervals. The figure contains results from LD genomic regions with confidence intervals band not including 0. Segment size indicates the number of SNPs in the implicated loci segment selected (largest segment if overlaps existed).

### Gene expression/eQTL analysis

We used COLOC [19] and expression data from The Genotype Tissue Expression (GTEx) project (v8) [20] to identify candidate causal genes at significant local genetic covariance segments between sU and eGFR. Forty-one of the 188 distinct significant shared loci (21.8%) are shown to modify the expression of 90 candidate causal genes colocalized with the covariance signals (S4 Table). Of note are 5 genes with covariance signals and colocalized eQTL that are expressed in the kidney: *FGF5, ARL6IP5, TRIM6, BCL2L14* in *cis*, and *NTRK1* in *trans*.

### Validation

We performed a validation analysis with the Atherosclerosis Risk in Communities Study (ARIC) utilizing 8,752 distantly related white subjects with 739,587 genotyped SNPs after standard quality controls on the phenotypes and genotypes. Some of the largest magnitude covariance estimates (*e*.*g*., *SHROOM3, SLC15A2*, and *SLC2A9*) were validated in terms of effect size, though they were not necessarily loci significant for local genetic covariance, likely due to the substantially smaller sample size in ARIC compared to the UK Biobank. Similar to the covariance estimates, the variance estimates were validated only in the largest effect size loci, such as *SHROOM3* and *GATM* for eGFR variance, and *SLC2A9* and *ABCG2* for sU.

## Discussion

The goal of this study was to infer the shared genetic architecture of sU (causal for gout), and eGFR (causal for CKD). Our results highlight genes that may be involved in the observed relationship between the traits. In this study, we utilized the large-scale UK Biobank and formal statistical inference from local Bayesian regression models to estimate local genetic covariances to identify shared loci. Our results demonstrated that genetic covariance between eGFR and sU was widespread across the genome. Our method identified 188 distinct LD segments with shared genetic effects between eGFR and sU, the majority of which agree with the global negative correlation directionality [11,12]. Many of the loci identified were previously only known to be associated with one of the two traits, demonstrating that the set of loci contributing to both traits is substantially larger than previously thought.

Out of the significant shared loci, almost all showed negative local genetic covariance estimates. This is consistent with the overall genetic covariance directionality [11,12], indicating that they either contribute to worsening kidney function (decreasing eGFR or higher sCr) and increasing sU, or *vice versa*. Interestingly, there were 10 significant shared loci with positive local genetic covariance estimates: *NRBP1*/*IFT172*/*FNDC4*/*GCKR, CPS1, SLC7A9*/*CEP89, A1CF, PIP5K1B, BCAS3, B4GALT1, OR52H1*/*HBG2*, and *LRP2*, which had 2 distinct positive covariance loci. Positive covariance indicates that the genomic region either contributes to increasing sU and improved kidney function, or decreasing sU and worsening kidney function. Two of the 10 loci with a positive signal, *GCKR* and *CPS1*, are mainly expressed in the liver and one, *LRP2*, is mainly expressed in the kidney [20]. Segments that have directionality opposite of the overall genetic correlation are masked by the overall correlation estimate, but our local method can distinguish them.

Segments encompassing the *SLC2A9* locus had some of the largest local genetic covariance estimates and showed both positive and negative estimates. Urate transporters *SLC2A9* and *ABCG2* have the largest GWAS effect sizes for sU, accounting for a 4-5% of variance in sU [11,21–24]. However, only one small magnitude segment in *SLC2A9* was significant for covariance. Interestingly, one SNP in that segment is rs16890979, which is a missense variant that has been identified in numerous sU GWAS [25–27]. *ABCG2* also had LD segments with both positive and negative estimates of large magnitude, but no segments from the *ABCG2* locus were significant for covariance. Our results demonstrate that, with the exception of one segment, segments in both *SLC2A9* and *ABCG2* loci are associated with just sU levels, but are not pleiotropic regions for sU and eGFR. A similar phenomenon is observed with the largest magnitude eGFR gene, *SHROOM3*. That is, none of the segments found in *SHROOM3* were significant for local genetic covariance. This exemplifies that the loci driving the genetic correlation between these two traits are not necessarily the loci found from analysing the traits individually.

Previous research investigating pleiotropic genetic loci between serum urate and eGFR has implicated loci as shared if signals of association obtained from marginal single-marker regressions (*e*.*g*., GWAS) for both traits are colocalized based [14]. Leask *et al*. [14] recently compared overlapping loci between two large GWAS, one of sU and the other kidney function [11,28], and found 35 independent colocalized loci. Our results validate 25 of these 35 loci, and all but 3 loci (*DACH1, CPS1*, and *INS-IGF2*) had covariance directionality that matched the directionality of effects found by Leask *et al*. [14]. The LBR method we utilized also identified numerous novel loci with significant local genetic covariance for sU and eGFR, including *LINC01101, KCNS3, CYP24A1, PAX8, ARL15, CHD9, IGF1R, PIP5K1B*, and *SLC7A9/CEP89*.

Our covariance approach has direct implications for assessing causal relationships between exposures using Mendelian randomization (MR). Pleiotropic genetic variants violate assumptions of univariate MR, however, they are useful in multivariable MR that can simultaneously assess the causal effects of multiple risk factors on an outcome [29]. For example, genetic variants from *SLC2A9* and *ABCG2* may be valid instrumental variables to use in MR to test for a causal effect of sU on CKD, however, the loci listed in Table 1 and S1 Table would not. In fact, *SLC22A11* has previously been identified as a pleiotropic variant that may improve kidney function through its activity in raising urate levels [23]. MR has previously been used to show that serum urate is not causal of CKD [30], however, Jordan *et al*. noted significant pleiotropy in the genetic variants used in their study, which they attempted to counter using MR techniques robust to pleiotropy. Of the 26 SNPs used by Jordan *et al*., two were identified by us as shared (gene indicated next to the SNP), and six imputed variants were located within one of our significant pleiotropic regions (these SNPs were not in our genotyping platform), between sU and eGFR: rs1260326 (*GCKR*), rs17050272 (*LINC01101*), rs729761, rs10480300, rs10821905, rs3741414, rs1394125, and rs6598541.

Our eQTL analysis of the segments significant for local genetic covariance uncovered numerous genes of interest, such as *SLC7A9*, which encodes a solute transporter largely expressed in the small intestine, *A1CF*, which encodes a protein involved in apolipoprotein B synthesis in the liver, and *TRIM6*, which encodes an E3 ubiquitin ligase involved in interferon gamma signalling and innate immune response with high expression levels in the kidney [20]. The genes uncovered from the eQTL analysis will be particularly interesting for future study, as they will likely aid our understanding of the relationship between kidney function and sU.

This study had several strengths. Through our novel statistical approach of obtaining genetic covariance estimates from conditional LBR models in very large datasets, we have uncovered numerous novel genomic regions that can be defined as shared genetic regions for sU and eGFR. The approach presented in this paper was applied in the context of sU and eGFR, but it could be applied to any pair of continuous traits. While local genetic correlation estimates can theoretically be obtained from fitting local multivariate mixed models that utilize genetic and phenotypic information on sU and kidney function, a limitation is that with increasingly large datasets this is computationally challenging. Our method overcomes this limitation by enabling us to obtain local genetic covariance point estimates genome wide while still utilizing the large size of the UK Biobank.

The local shared genomic regions we have uncovered in this study can provide insight into the relationship between hyperuricemia and CKD, elucidating the biological mechanisms underlying the traits. This will help to further understanding of the genetic basis of hyperuricemia and CKD.

## Materials and Methods

This study used 333,542 Caucasian unrelated subjects from the UK Biobank. Subjects missing phenotypes of interest for both of their two visits were excluded from the analysis. The UK Biobank used the custom UK Biobank Axiom™ Array by Affymetrix to genotype study participants [31]. Quality control involved removing SNPs that had a minor allele frequency less than 1% or a missing call rate greater than 5%, resulting in 607,490 autosomal chromosome (1-22) SNPs [32].

### Identification of unrelated samples

We used the R package BGData [33] to compute the expected proportion of allele sharing among UK Biobank individuals with the additive genomic relationship matrix ***G***, 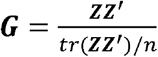, where ***Z*** is a matrix of centered genotypes. That is, ***Z***_*ij*_ = *x*_*ij*_ -2*p*_*j*_ where *x*_*ij*_ is the number of copies of the reference allele at the *j*^th^ loci of the *i*^th^ individual and *p*_*j*_ is the frequency of the reference allele of the *j*^th^ loci. In a homogeneous sample, *gij* (where *i*≠ *j*) can be considered as an estimate of the relatedness between subjects *i* and *j*. If *g*_*ij*_ ≥ 0.1 they were excluded from the sample.

### Phenotypes

sU and sCr data was obtained from the first visit. For the small number of participants (0.28%) that did not have phenotype data of interest collected at the first visit, we retrieved data from the second visit.

eGFR is an indicator of renal function and used to ascertain CKD. In this study, we defined eGFR using the abbreviated Modification of Diet in Renal Disease (MDRD) equation, which uses fewer variables than others yet performs just as well [34], with a modification to include a calibration factor to correct for the variability of sCr measures across laboratories and time [35]: *eGFR* = 186.3 × (*sCr* – 0.24) ^-1.154^ × *Age* ^-0.203^ × (0.742 if *Female*).

For both eGFR and sU, we took a *log* transformation to normalize their distributions and preadjusted by age, sex, and the first 5 SNP-derived principal components using ordinary least squares.

### LBR model specification

Following Funkhouser *et al*. [17], we fit a series of LBR models based on a core chunk of 10,000 contiguous SNPs, and an overlapping flanking buffer of 500 SNPs taking the form of 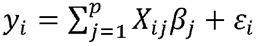 for *p*=11,000 (core SNPs plus two flanking buffers). This method will be a robust substitute for a single whole-genome regression since LD spans over relatively short regions of SNPs in the UK Biobank Axiom™ Array and a homogeneous unstructured sample like the one used here [17]. Each LBR utilizes the BGLR R package [18] with a BayesC prior for the SNP effects, which has a point mass at zero and Gaussian slab. This prior performs variable selection to zero-out some SNPs and reduce the number of SNPs entering the model [36]. The Markov chain Monte Carlo algorithm for BayesC involves a Gibbs-sampler sequence of steps with the full-conditional posterior distributions [36]. The Markov chain Monte Carlo runs had long chains of 75,000 iterations, with a burn-in of 2,000 samples and thin of 5 that were discarded.

### LBR implementation

The LBR had the following phenotypes as the response variable: sU (*y*_*1*_), eGFR (*y*_*2*_), and sU + eGFR (*y*_*3*_). For each of these three response models, we ran the LBR models across the genome, and obtained genetic variance estimates within each LD segment. That is, for each *y*_*i*_ model, we estimated the genetic variance for each LD segment as 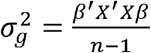 using only the SNPs within each LD segment. This allows us to then obtain a point estimate for the genetic covariance within each LD segment, we leveraged the fact that 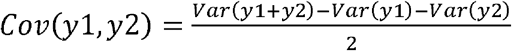.

Since each variance component comes from a separate series of LBR models, even though we have interval estimates for each of the three variance components, we cannot directly obtain an interval estimate for the covariance between sU and eGFR since there is not a closed form solution for the standard error estimates for the genetic covariance estimates. Therefore, we obtained interval estimates for select LD segments with a resampling method (described below). A visual summary of the local covariance pipeline can be found in S1 Fig.

### Defining local, LD-based segments

Local genetic covariance inference from an individual SNP is problematic due to underlying LD structures, so we identified SNP regions in strong LD, and obtained variance estimates based on these LD segments of SNPs rather than using single individual SNPs [17]. We used an overlapping sliding technique to obtain these local LD segments [16,17]. For each seed SNP ***x***_*j*_, we sequentially identify SNPs in both directions (***x***_*j\**_) surrounding the seed SNP and include them in segment *j* if Corr(***x***_*j*_, ***x***_*j\**_) ≥ 0.1. In a simplified example, if SNP ***x***_*j*_ has adequate pairwise correlation with 2 SNPs to the left, and 1 SNP to the right, the segment for that SNP would be defined as the set of SNPs: {***x***_*j*-2_, ***x***_*j*-1_, ***x***_*j*_, ***x***_*j*+1_}. That is, Corr(***x***_*j*_, ***x***_*j*-1_) ≥ 0.1 and Corr(***x***_*j*_, ***x***_*j*-2_) ≥ 0.1 and Corr(***x***_*j*_, ***x***_*j*+1_) ≥ 0.1. However, our algorithm also involved an allowance for one SNP in the sequential process to not meet this correlation criterion, to allow for a brief loss of LD or minor mapping errors, and the SNP was still included in the LD segment. Continuing with the previous example, even if Corr(***x***_*j*_, ***x***_*j*-1_) < 0.1, if Corr(***x***_*j*_, ***x***_*j*-2_) ≥ 0.1, then the set would still include both ***x***_*j*-2_ and ***x***_*j*-1_. The LD block ends when two SNPs sequentially did not meet the criteria described above.

### Confidence interval estimates of the local covariances

We estimated CIs for the most interesting LD regions based on bootstrapping methods. Because of the computational demands required by bootstrap resampling techniques with very large sample sizes, we preselected peaks to limit the CI estimates only to regions of interests. We considered GWAS significant variants for sU and CKD markers (sCr and eGFR) as indicators of loci considered regions of interest, so we applied a 100-SNP buffer to each side of each GWAS locus. All LBR regions of the SNPs of interest plus contiguous flanking SNPs were included in the model. The LBR models were identical to the description above (LBR model specification). We ran 200 bootstrap replicates using a sample of size n=333,542 with replacement for each response model and averaged the iterations to obtain bootstrap covariance estimates. We obtained the 2.5% and 97.5% quantiles from the iterations to obtain 95% CIs for the bootstrap covariance estimates (Table 1 and S1 Table).

### Multiple testing adjustment

Statistical significance was also conservatively estimated based on a Bonferroni multiple testing correction. We obtained normality-based p-values from a T-statistic from our bootstrapped covariance estimates and divided that by the standard error estimates obtained from the standard deviation of the bootstrap iterations. This value was then compared to the standard normal distribution. We performed this on 14,802 LD regions, which determined the conservative Bonferroni multiple testing adjustment.

### Genome-Wide Association Studies

GWAS were performed to identify SNPs significantly associated with sU and sCr using single marker linear regression models in the UK Biobank sample. Each GWAS was performed for *k*=607,490 SNPs that passed quality control (described above). The sU GWAS used a sample size of n=288,831 unrelated, white participants. Participants were excluded from the sU GWAS if they were missing the sU phenotype, if they were not between the ages of 40 and 69 years old, if their genotypes did not pass quality control, and if they had a primary or secondary diagnosis of kidney disease. The sCr GWAS used a sample size of n=301,594 distantly related, white participants. Participants were excluded from the sCr GWAS if they were missing the sCr phenotype. Both GWAS were performed using the following model for each SNP variant *j*={1, …, *k*}: **y** = **µ** + **Xβ** +**W**_**i**_**s**_**i**_ + ε. where **y**=(y, …, y)*’* is the vector of phenotypic observations for sU, eGFR, or sCr, **µ** is a vector of the overall mean, and **X** is a design matrix connecting the fixed effect levels to the observations. The fixed effects included sex, age, and the first 5 SNP-derived principal components. Additionally, β is a vector with the corresponding effects, **W**_**i**_ is aε _1 n_ vector with the *j*^th^ SNP, **s**_**i**_ is the additive genetic effect of the *j*^th^ SNP, and ε =(ε_1_, …,ε_n_)*’* is the vector of the residuals. A variant was considered significant if it had a p-value < 5e-8. The GWAS summary statistics can be found in S2 Table.

### Gene expression/eQTL analysis

A colocalization analysis was performed between GWAS significant markers for sU and sCr and the publicly available eQTL data from GTEx v8 [19]. The R package COLOC was used, which implements a Bayesian test that analyses a single genomic region and identifies LD patterns in that locus using SNP summary statistics and the associated minor allele frequencies. The lead variant for both sCr and sU was used at each significant covariance segment with a surrounding 500 kb buffer in the GTEx database. The Contextualizing Developmental SNPs using 3D Information algorithm [37,38] was modified to identify long-distance regulatory relationships for the lead sU and sCr variants at each significant covariance segment within a 500 kb window. eQTL data for variants +/-500 kb of the lead variant were also extracted from GTEx and then COLOC was used to assess if the significant *cis*-and *trans*-eQTL identified were colocalized with sCr and sU signals. The eQTL was required to have a posterior probability of causality (PPC) of at least 0.5 for both traits, along with a PPC of at least 0.8 for one of the two traits.

## Data Availability

**Data**
No primary data was generated in the project. Our project utilizes existing resources already generated from the database of Genotypes and Phenotypes and the UK Biobank.
**Software**
All software produced in this project will be distributed as open source at R CRAN and at GitHub. Our research group's releases are extensively documented. Our QuantGen GitHub page has numerous examples of our well-documented published software (https://quantgen.github.io/).

## Supporting information captions

**S1 Fig**. A visual summary of our methodology to obtain covariance estimates between sU and eGFR.

**S1 Table**. The effect sizes with corresponding confidence intervals, significance, the directionality of the association, and validations for all significant covariance regions.

**S2 Table**. The results from a UK Biobank GWAS in sU and sCr.

**S3 Table**. The 188 distinct loci implicated by the LD segments significant for genetic covariance, with eQTLs.

**S4 Table**. The eQTLs that colocalized with the sU and eGFR covariance signals.

## Acknowledgments

This study was funded by the National Institute of Arthritis and Musculoskeletal and Skin Diseases P50AR060772 (Insight CORT), and by Michigan State University. The Contextualizing Developmental SNPs using 3D Information algorithm was run on computing resources provided by the University of Auckland. JOS and TF were funded by the Dines Family Charitable Trust and HRC Explorer Grant (HRC 19/774). We would like to thank Michigan State University’s High Performance Computing Cluster for providing all additional computing resources.

