## Supplementary figures and images for "Local genetic covariance between serum urate and kidney function obtained from local Bayesian regressions"

### S1 Fig

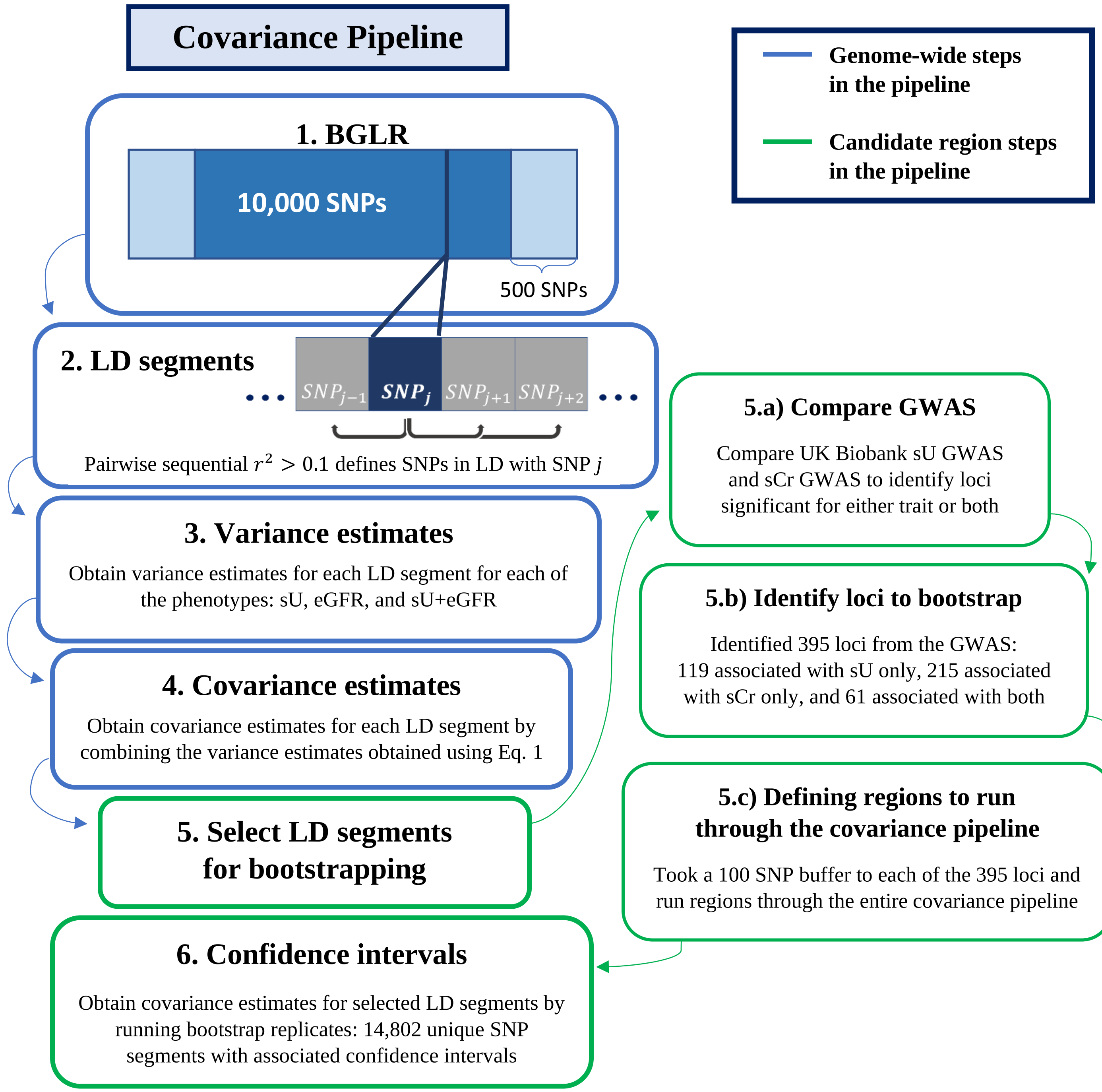
